# A global survey on changes in the supply, price and use of illicit drugs and alcohol, and related complications during the 2020 COVID-19 pandemic

**DOI:** 10.1101/2020.07.16.20155341

**Authors:** Ali Farhoudian, Seyed Ramin Radfar, Hossein Mohaddes Ardabili, Parnian Rafei, Mohsen Ebrahimi, Arash Khojasteh Zonoozi, Cornelis A J De Jong, Mehrnoosh Vahidi, Masud Yunesian, Christos Kouimtsidis, Shalini Arunogiri, Helena Hansen, Kathleen T Brady, Marc N Potenza, ISAM-PPIG Global Survey Consortium, Alexander Mario Baldacchino, Hamed Ekhtiari

## Abstract

**Background and aims:** COVID-19 has infected more than 13 million people worldwide and impacted the lives of many more, with a particularly devastating impact on vulnerable populations, including people with substance use disorders (SUDs). Quarantines, travel bans, regulatory changes, social distancing and ‘lockdown’ measures have affected drug and alcohol supply chains and subsequently their availability, price and use patterns, with possible downstream effects on presentations of SUDs and demand for treatment. Given the lack of multicentric epidemiologic studies, we conducted a rapid global survey within the International Society of Addiction Medicine (ISAM) network in order to understand the status of substance-use patterns during the current pandemic.

**Design:** Cross-sectional survey.

**Setting:** Worldwide.

**Participants:** Starting on April 4^th^, 2020 during a 5-week period, the survey received 185 responses from 77 countries.

**Measurements:** To assess addiction medicine professionals’ perceived changes in drug and alcohol supply, price, use pattern and related complications during the COVID-19 pandemic.

**Findings:** Participants reported (among who answered “decreased” or “increased”, percentage of those who were in majority is reported in the parenthesis) a decrease in drug supply (69.0%), and at the same time an increase in price (95.3%) globally. With respect to changes in use patterns, an increase in alcohol (71.7%), cannabis (63.0%), prescription opioids (70.9%), and sedative/hypnotics (84.6%) use was reported while the use of amphetamines (59.7%), cocaine (67.5%), and opiates (58.2%) was reported to decrease overall.

**Conclusions:** The global report on changes in the availability, use patterns and complications of alcohol and drugs during the COVID-19 pandemic should be considered in making new policies and in developing mitigating measures and guidelines during the current pandemic (and probable future ones) in order to minimize risks to SUDs.

**Competing interest:** Authors claimed no competing interest

## 1. Introduction

As of July 14, the COVID-19 pandemic has around 13 million cases of infection in more than 200 countries with above 570,000 overall deaths (1). Approximately 6 months after cases were first diagnosed, there remain few reliable treatments and no vaccines available and an increasing number of countries are experiencing dangerous COVID-19 transmission (2, 3). Among vulnerable populations to infection and its complications are people with substance use disorders (SUDs) (4). Both comorbid medical conditions in SUDs (such as cardiopulmonary diseases and related risk factors) and drug-drug interactions (between COVID-19 medications and abused substances or SUD treatment medications), along with other factors, may lead to people with SUDs experiencing more complications when encountering COVID-19 infections (4-6).

People with SUDs are vulnerable given marginalization, stigmatization, and poor access to health and social services (7, 8). According to risky behaviors and disadvantaged environments associated with SUDs, people with SUD may not only bear additional risks for COVID-19 but also experience poorer outcomes (4). Therefore, during the pandemic, gathering current information on the status of SUD is critical to support planning and mobilizing timely responses to minimize risks (4).

Alterations in alcohol and drug supplies may change prices and availability and therefore use patterns. The COVID-19 pandemic has resulted in quarantines, travel bans, regulatory changes and social distancing ‘lockdown’ measures globally, with impacts on supply chains. In the setting of COVID-19-related stressors, there may be decreases in drug and alcohol availability, increases in price and use patterns, and possible downstream effects on SUD presentations and treatment demands. Such changes could directly/indirectly affect people with SUDs and give rise to new challenges and additional needs in the field of addiction medicine. Drug shortages, as the United Nation Office for Drug and Crime (UNODC) reports, could have negative health consequences regarding transitioning to consumption of harmful domestically produced substances along with more dangerous patterns of drug use including shifting to injections and using shared drug administration equipment, especially in the case of heroin (9).

Additionally, the lack of drug supply may result in higher prices for some substances and bring financial burden to drug users and increase the odds of risky/illegal behaviors (4). Concurrently, as legal liquor shops may remain closed during the lockdown in some countries, multiple problems may occur ranging from alcohol withdrawal to toxicity and death due to shifting to low-quality homemade liquor and accidental methanol ingestion (4, 10).

People with SUDs could be exposed to some indirect risks during the COVID-19 era as well (5). For instance, as healthcare facilities become more difficult to access during lockdowns, people with SUDs may experience more difficulties relating to poor access to treatment centers. Socioeconomically disadvantaged backgrounds and diminished availability of public transportation may exacerbate such concerns (4, 5, 11), especially for individuals receiving daily prescriptions of opioid substitution therapy (4). Professional authorities and health policy makers are expected to proactively address such emerging needs. However, lack of reliable data complicates the generation and implementation of evidence-based policies.

Although some activities and reports from different worldwide organizations have initially responded to the COVID-19 pandemic, data provided have been limited and, in some occasions, as UNODC has reported, the information base for analyses has been restricted and feasibility of implementation unknown (12-17). Thus, a vacancy exists for a comprehensive report describing the global situation with respect to drug use, drug supply and related complications.

In order to formulate a comprehensive health response, it is important to understand alcohol and drug markets (availability and price), use patterns and related complications and how they may have changed during the pandemic. Designing a global in-depth epidemiologic study, apart from questions about its feasibility, is challenging during the pandemic. Therefore, the International Society of Addiction Medicine (ISAM) designed a comprehensive global survey and collected expert opinions from their membership on perceived changes on substance use situation and health system responses around the first week of April 2020 in what aims to be a longitudinal study (18).

Here, we report results from the first round of the ISAM global survey on drug and alcohol use, price, supply and complications during the COVID-19 pandemic. We hypothesized that drug and alcohol use would increase, prices would increase, supply would decrease, and complications would increase and that results would differ by region (given differential spread of COVID-19 and regional responses to the COVID-19 pandemic). We hope current data will help to address the urgent need for more accurate information about the status of drug and alcohol use in the current pandemic and provide information about appropriate modifications in health system services to respond to the emerging demands in the current pandemic and similar potential pandemics in the future.

## 2. Methods

### 2.1. Sample

The complete study protocol has been previously published (18). The ISAM email list (and subsequent snowballing methodologies) of addiction medicine professionals across the world (n= 2287) were contacted on April 4^th^, 2020 by email with an invitation to participate in the study by clicking on a link to the online survey. The invitees were informed that the survey will ask about their ‘opinions and information towards COVID-19 pandemic impact on SUDs. Data collection was concluded on May 8^th^, 2020.

### 2.2 Questionnaire

The questionnaire consisted of 92 questions in two main sections: (1) situational assessment during the pandemic and (2) health response to the pandemic. This paper provides an analysis of data obtained from the situation assessment section of the survey concerning changes in drug use, supply, price, risky behaviors, as well as related measures namely morbidities, mortalities and overdose rates during the COVID-19 pandemic period in different countries (18). Questions on the situational assessment section of the survey are available in supplementary methods.

### 2.3 Statistical Analysis

All statistical analyses were conducted using RStudio (v. 1.2.1335). Descriptive data are presented as means and percentages for each country’s response, as well as the average of the global responses.

### 2.4 Ethics Approval

The survey protocols and all materials, including the survey questionnaires, received approval from the University of Social Welfare and Rehabilitation Sciences, ethics committee in Tehran, Iran (Code: IR.USWR.REC.1399.061).

## 3. Results

### 3.1. Responders’ global distribution

Overall, 185 responders from 77 countries participated. Eight responses were excluded because of insufficient information provided (The “insufficient information” was predetermined as having more than 50% of ‘I do not know’ responses). Data from 177 responders were analyzed. The list of the countries that provided information for this survey is available as a supplement (Supplementary Method 1). Figure 1 depicts a map of responders’ global distribution.

**Figure 1.**
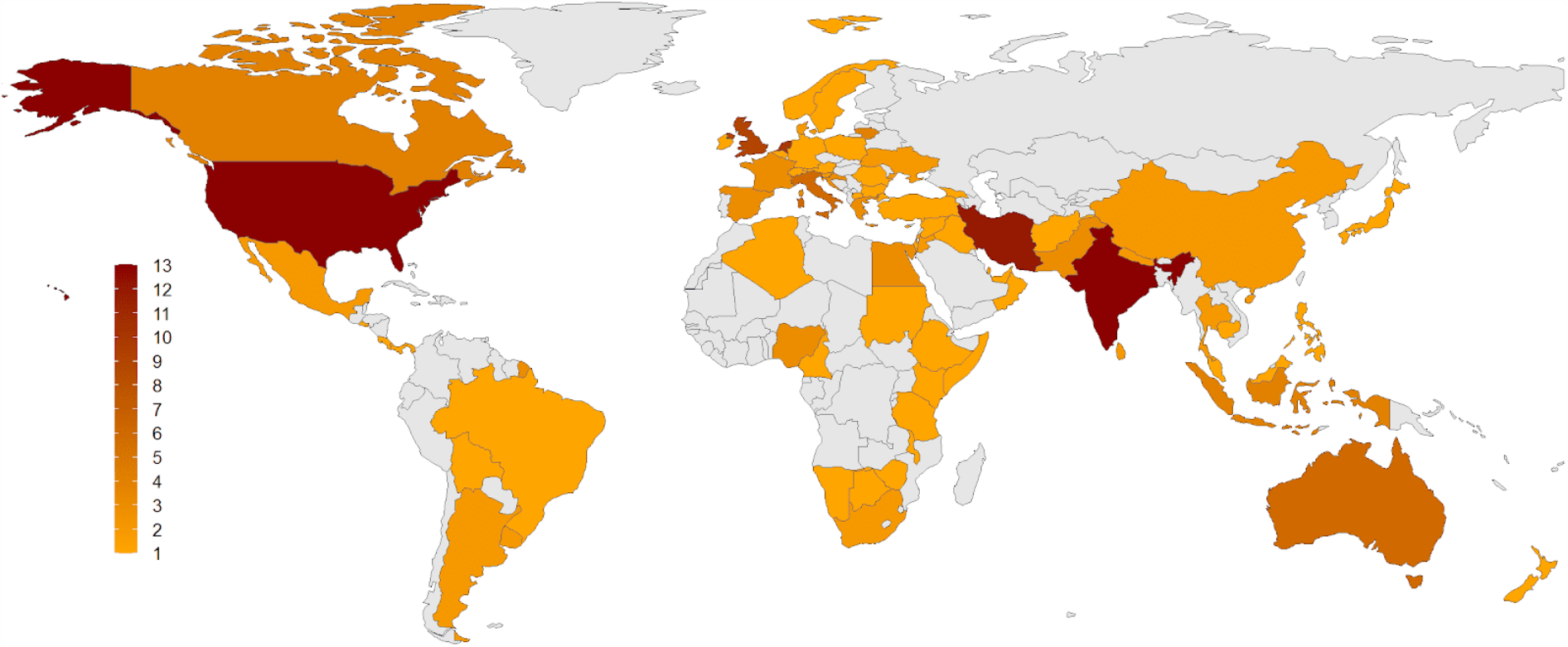
Global distribution of the responders to the survey. The survey involves 177 responders from 77 countries around the world, ranging from 1 to 13 participants from each country as demonstrated as a color spectrum from orange to dark red.

### 3.2. Responders’ demographic characteristics

Responders consisted of 111 males (62.7%), 62 females (35%), and 4 people (2.3%) who selected “other” or preferred not to disclose their gender. The mean age of responders was 46.51 ± 10.78 years. Most responders were medical professional (MDs) (n= 148, 83.6%), and the most frequent primary discipline was psychiatry (n=95, 53.7%). Information related to responders’ main disciplines and academic degrees is shown (table 1).

**Table 1.**
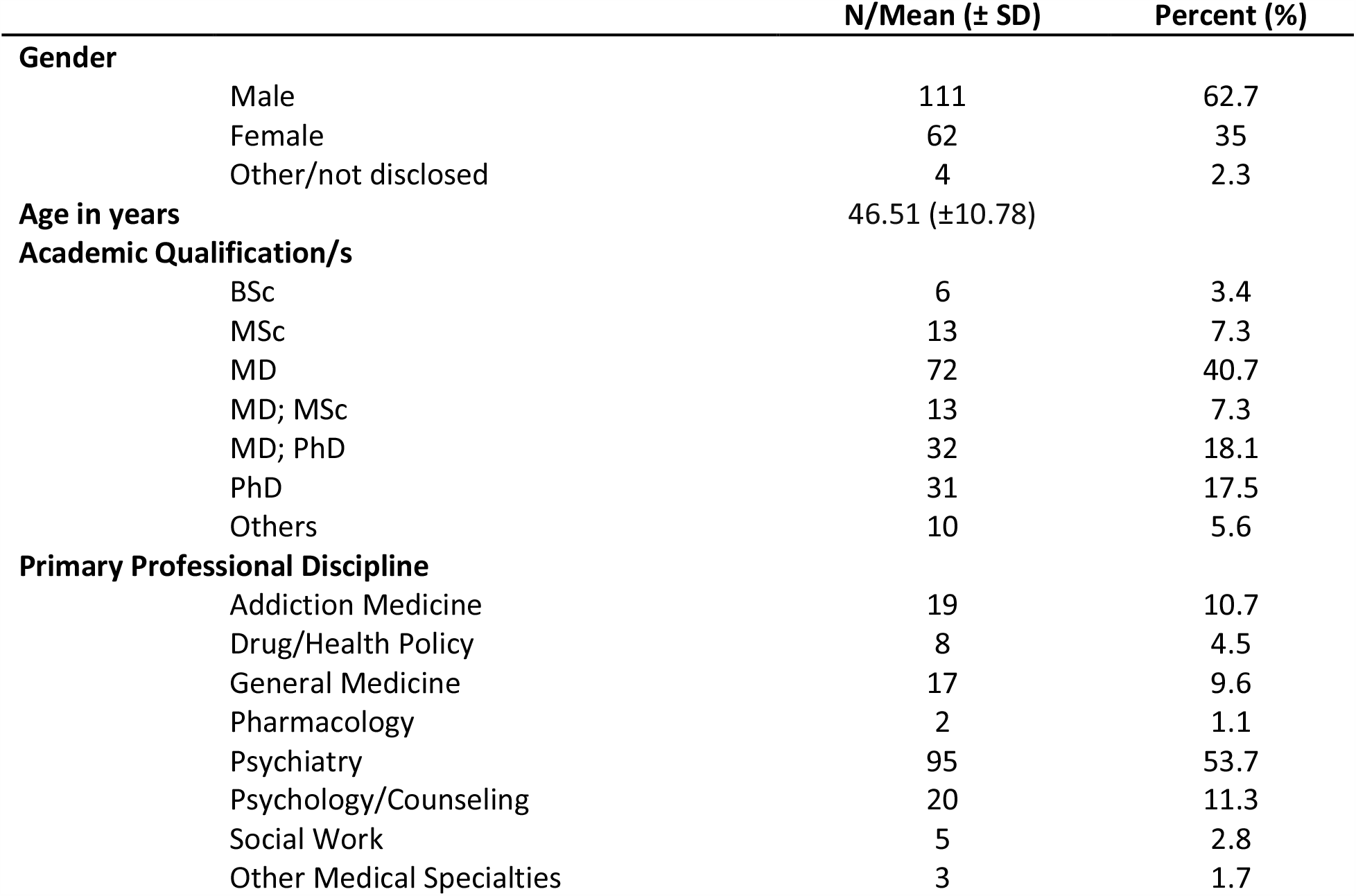
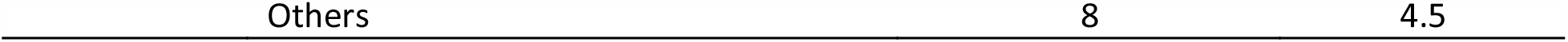
The demographic and professional information of survey responders including their gender, age, academic degree and primary discipline.

### 3.3. Drug use during Pandemic

Responders provided information about drug use changes in their countries during the COVID-19 pandemic. Over 63% (n=49), 42% (n=32), 64% (n=50) and 41% (n=32) of countries reported that use of alcohol, cannabis, sedatives and prescription opioids increased, respectively. Conversely, opiates, amphetamine and cocaine use have seen a decrement in 31% (n=24), 29% (n=22), and 29% (n=23) of the countries, respectively. Perceived drug use changes by country is shown (figure 2, table 2). Details of drug use changes are reported in supplementary materials.

**Table 2.** Summary of the survey responses in different sections related to situational assessment including responders’ information about changes in alcohol and drug use pattern, supply, price, morbidity and mortality and overdose.

|  | Responders (177) |  |  |  |  |  |  |  | Countries (77) |  |  |  |  |  |  |  |
| --- | --- | --- | --- | --- | --- | --- | --- | --- | --- | --- | --- | --- | --- | --- | --- | --- |
|  | Decrease |  | No Change |  | Increase |  | Others |  | Decrease |  | No Change |  | Increase |  | Others |  |
|  | N | % | N | % | N | % | N | % | N | % | N | % | N | % | N | % |
| **Use** |  |  |  |  |  |  |  |  |  |  |  |  |  |  |  |  |
| Alcohol | 43 | 24% | 13 | 7% | **109** | **62%** | 12 | 7% | 19 | 25% | 6 | 8% | **49** | **63%** | 3 | 4% |
| Cannabis | 44 | 25% | 35 | 20% | **75** | **42%** | 23 | 13% | 20 | 26% | 17 | 22% | **32** | **42%** | 8 | 10% |
| Opiates | **53** | **30%** | 43 | 24% | 38 | 21% | 43 | 24% | **24** | **31%** | 16 | 20% | 14 | 18% | 23 | 30% |
| Amphetamines | **49** | **28%** | 30 | 17% | 33 | 19% | 65 | 37% | **22** | **29%** | 15 | 20% | 14 | 18% | 26 | 33% |
| Cocaine | **52** | **29%** | 28 | 16% | 25 | 14% | 72 | 41% | **23** | **29%** | 15 | 19% | 10 | 14% | 29 | 38% |
| Sedatives | 19 | 11% | 24 | 14% | **105** | **59%** | 29 | 16% | 5 | 6% | 9 | 11% | **50** | **64%** | 14 | 18% |
| Presc. Opioids | 27 | 15% | 35 | 20% | **66** | **37%** | 49 | 28% | 8 | 11% | 16 | 21% | **32** | **41%** | 21 | 27% |
| **Supply** |  |  |  |  |  |  |  |  |  |  |  |  |  |  |  |  |
| Alcohol | **62** | **35%** | 49 | 28% | 52 | 29% | 14 | 8% | **26** | **34%** | 21 | 28% | 24 | 31% | 6 | 7% |
| Cannabis | **62** | **35%** | 46 | 26% | 33 | 19% | 36 | 20% | **29** | **37%** | 18 | 24% | 15 | 20% | 15 | 19% |
| Opiates | **71** | **40%** | 34 | 19% | 18 | 10% | 54 | 31% | **31** | **41%** | 14 | 18% | 6 | 8% | 26 | 33% |
| Amphetamines | **54** | **31%** | 34 | 19% | 19 | 11% | 70 | 40% | **29** | **38%** | 14 | 18% | 7 | 9% | 27 | 35% |
| Cocaine | **56** | **32%** | 32 | 18% | 15 | 8% | 74 | 42% | **26** | **34%** | 14 | 18% | 7 | 9% | 30 | 39% |
| **Price** |  |  |  |  |  |  |  |  |  |  |  |  |  |  |  |  |
| Alcohol | 5 | 3% | **91** | **51%** | 57 | 32% | 24 | 14% | 3 | 4% | **42** | **54%** | 23 | 29% | 10 | 13% |
| Cannabis | 4 | 2% | 51 | 29% | **70** | **40%** | 52 | 29% | 2 | 3% | 23 | 30% | **30** | **39%** | 22 | 28% |
| Opiates | 2 | 1% | 33 | 19% | **75** | **42%** | 67 | 38% | 2 | 2% | 14 | 18% | **29** | **37%** | 33 | 43% |
| Amphetamines | 3 | 2% | 33 | 19% | **57** | **32%** | 84 | 47% | 2 | 3% | 13 | 17% | **26** | **34%** | 35 | 46% |
| Cocaine | 1 | 1% | 35 | 20% | **51** | **29%** | 90 | 51% | 0 | 0% | 18 | 23% | **21** | **28%** | 37 | 49% |
| **Morbidity and mortality** |  |  |  |  |  |  |  |  |  |  |  |  |  |  |  |  |
| Alcohol | 7 | 4% | 34 | 19% | **72** | **41%** | 64 | 36% | 5 | 7% | 16 | 21% | **27** | **35%** | 29 | 38% |
| Drug | 7 | 4% | 34 | 19% | **68** | **38%** | 68 | 38% | 5 | 6% | 14 | 18% | **28** | **36%** | 30 | 39% |
| **Overdose** | 14 | 8% | **53** | **30%** | 35 | 20% | 75 | 42% | 7 | 9% | **24** | **32%** | 14 | 18% | 32 | 42% |

**Figure 2.**
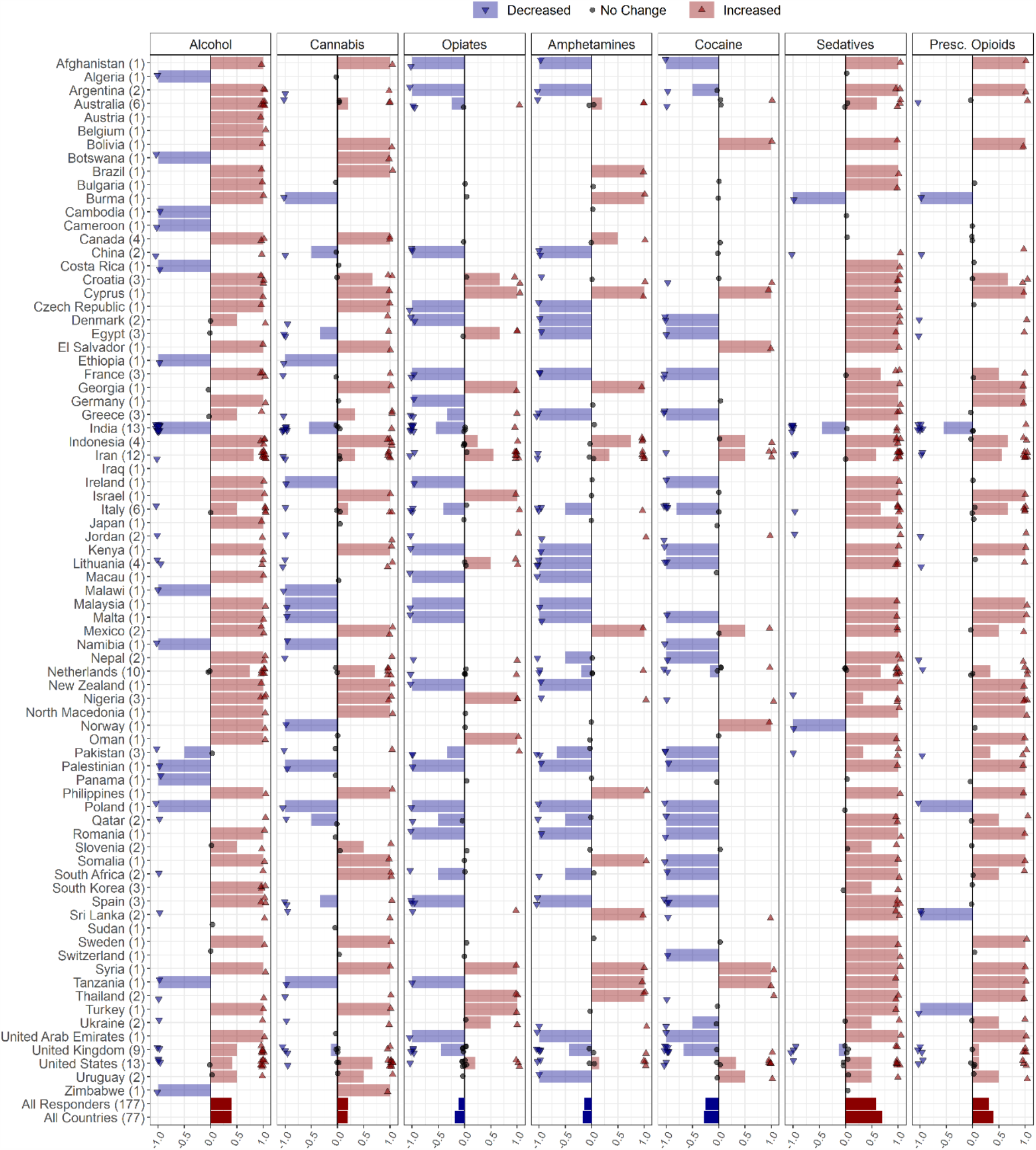
Changes in alcohol and drug use during the COVID-19 pandemic reported by 177 responders from 77 countries globally. Responders were asked to report changes in alcohol, amphetamines, cannabis, cocaine, opiates, prescribed opioids and sedative-hypnotics use with the following options: *Increased, Decreased, Not changed, I do not know, Number of users is very low/none*. Countries’ names are sorted in alphabetical order and the number of responders from each country is in parenthesis following the country name. Each response is indicated as a single dot for *no change* or up and down triangles for *increased* and *decreased* answers respectively, with a minor jitter for better visualization. The reported answers are represented as -1 for *decreased*, 1 for *increased* and 0 for *no change. I do not know* and *Number of users is very low/none* answers are not shown in the figure. The mean of all responses, regardless of their originated countries and without considering those who didn’t know the answer or reported very low/none number of users, alongside the average answers of all countries, regardless of the number of responders in each country, are addressed in the last two rows below the countries’ names. (Pres. Opioids: prescription opioids).

Responders were also asked to report changes in behavioral addictions (gaming/gambling) in their countries through the following options: Increased, Decreased, No change, I do not know. 85.7% (n=66) of the countries reported that behavioral addictions rates had increased, whereas 14% (n=11) of the countries reported that behavioral addictions rates had decreased in their countries during the COVID-19 pandemic (Supplementary Figure 1.).

### 3.4. Drug Supply

Responders provided information about perceived drug supply changes in their countries during the COVID-19 pandemic. The drug categories included: alcoholic beverages, cannabis (including marijuana and synthetic cannabinoids such as spice, K2, etc.), opiates (including opium, heroin, opium residue, etc.), amphetamine-type stimulants (including amphetamine, methamphetamine, MDMA, etc.), and cocaine (including crack cocaine).

Decreased supply patterns for all substances were noted. A decrement was reported in supply in 34% (n=26) of the countries for alcohol, 37% (n=29) for cannabis, 41% (n=31) for opiates, 38% (n=29) for amphetamines, and 24% (n=26) for cocaine (Figure 3, Table 2). Details of drug supply changes are reported in supplementary materials.

**Figure 3.**
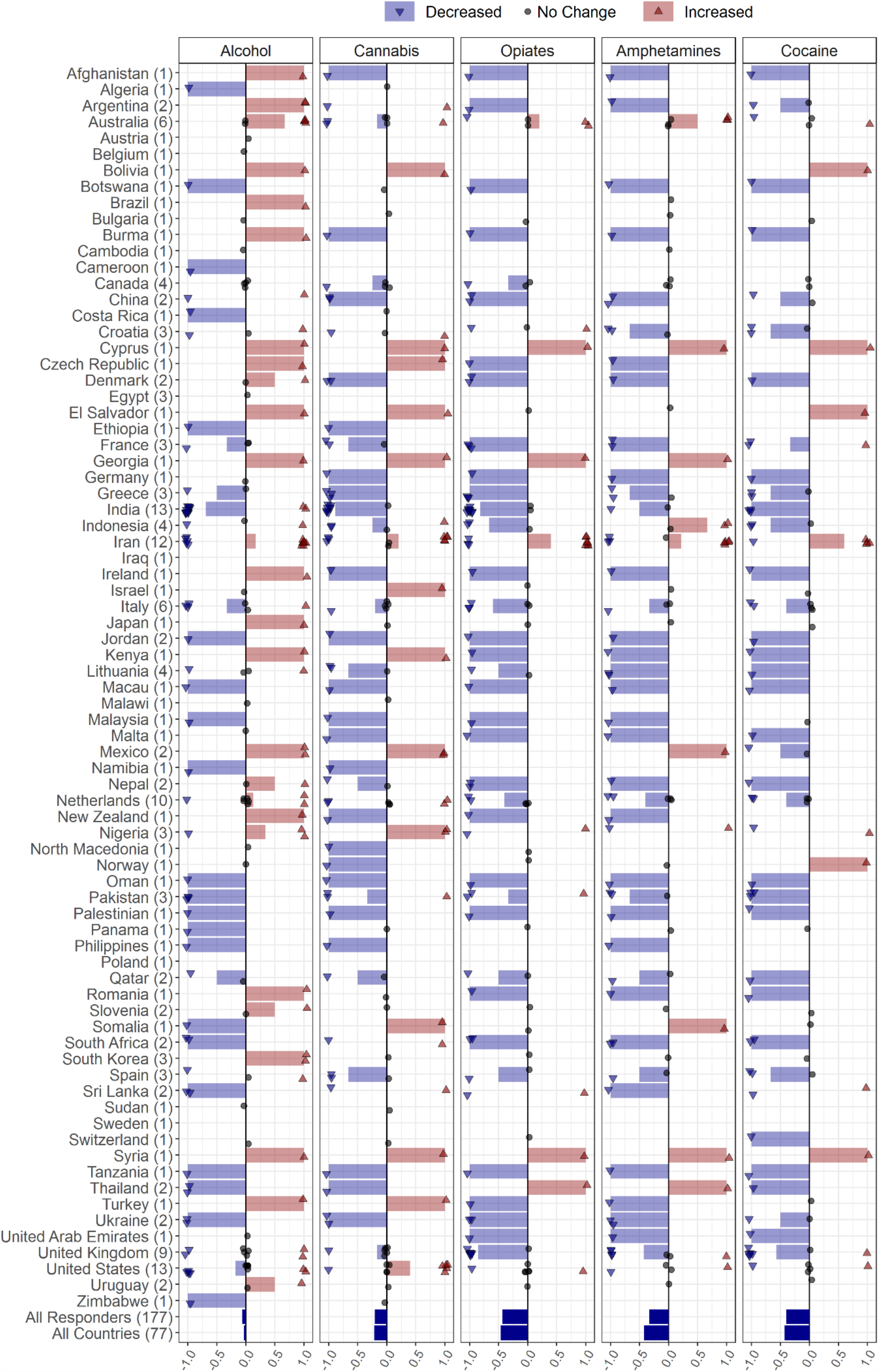
Changes in alcohol and drug supply during the COVID-19 pandemic reported by 177 responders from 77 countries globally. Responders were asked to report changes in supply of alcohol, amphetamines, cannabis, cocaine and opiates through the following options: *Increased supply, decreased supply, no change, I do not know*. Countries’ names are sorted in alphabetical order and the number of responders from each country is in parenthesis following the country name. Each response is indicated as a single dot for *no change* or up and down triangles for *increased* and *decreased* answers respectively, with a minor jitter for better visualization. The reported answers are represented as -1 for *decreased*, 1 for *increased* and 0 for *no change; I do not know* answers are not shown. The mean of all responses, regardless of their originated countries and without considering those who didn’t know the answer, alongside the average answers of all countries, regardless of the number of responders in each country, are addressed in the last two rows below the countries’ names.

### 3.5. Drug price

Responders provided information regarding perceived drug price changes in their countries during the COVID-19 pandemic. The price of cannabis, opiates, amphetamines and cocaine increased in 39% (n=30), 37% (n=29), 34% (n=26), and 28% (n=21) of the countries, respectively. Alcohol price was reported as unchanged in 54% (n=42) of countries (Figure 4, Table 2). Details of drug price changes are reported in supplementary materials.

**Figure 4.**
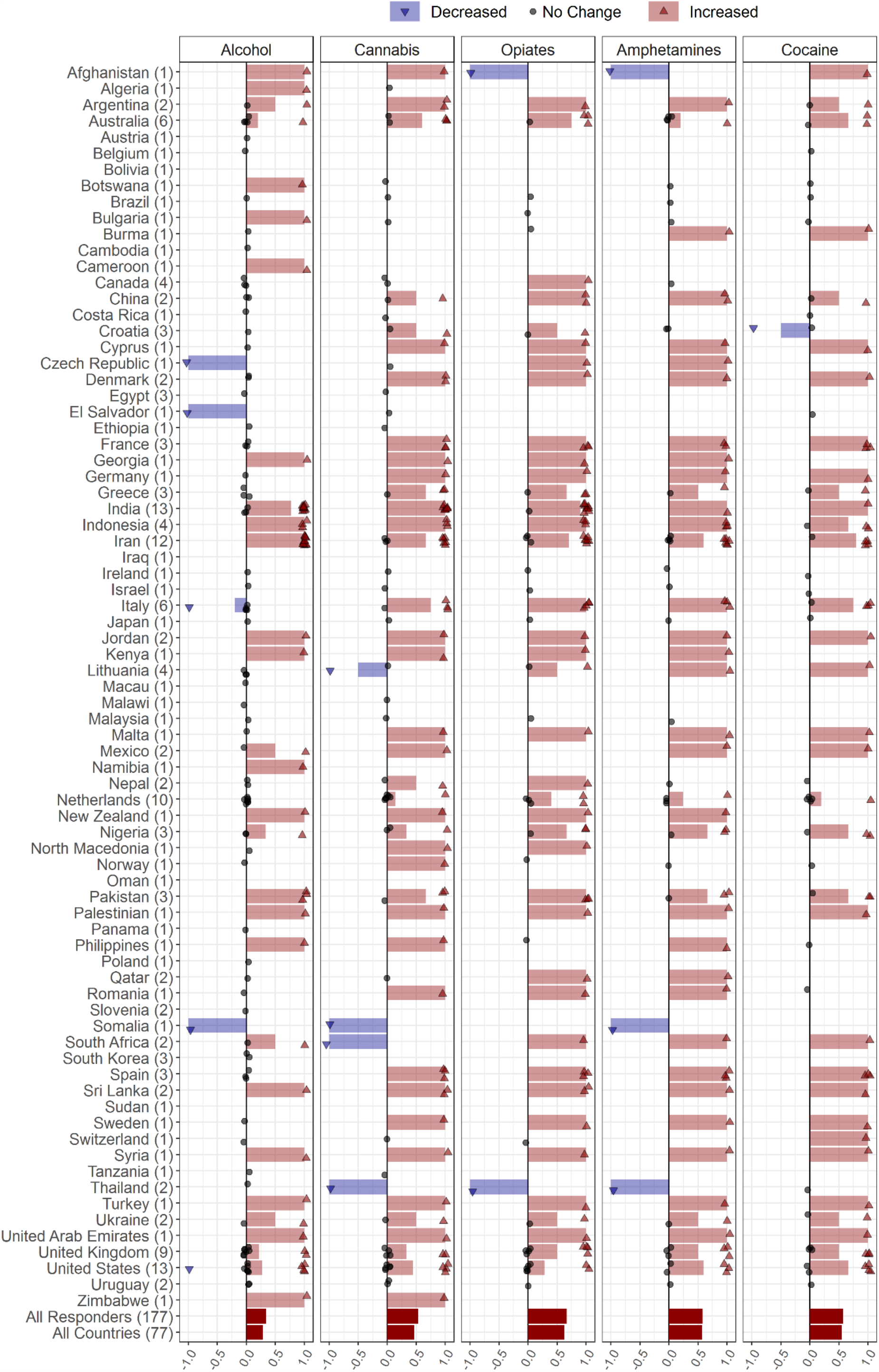
Changes in alcohol and drug prices during the COVID-19 pandemic reported by 177 responders from 77 countries globally. Responders were asked to report changes in alcohol, amphetamines, cannabis and opiates prices through the following options: *Price increased, Price decreased, Price did not change, I do not know*. Countries’ names are sorted in an alphabetic order and the number of responders from each country is in parenthesis following the country name. Each response is indicated as a single dot for *no change* or up and down triangles for *increased* and *decreased* answers respectively, with a minor jitter for better visualization. Reported answers are represented as -1 for *decreased*, 1 for *increased* and 0 for *no change; I do not know* answers are not shown in the figure. The mean of all responses, regardless of their originated countries and without considering those who didn’t know the answer, alongside the average answers of all countries, regardless of the number of responders in each country, are addressed in the last two rows below the countries’ names

The information related to changes in drug price among different countries is shown in figure 4 and table 2.

### 3.6. Perceived morbidity and mortality (including overdose)

Responders provided information about whether morbidity and mortality, including fatal and non-fatal overdose rates, in their countries had changed during the COVID-19 pandemic. Mortality rates in people with alcohol use disorders (AUDs) and SUDs were reported to have increased in 35% (n=27) and 36% (n=28) of countries, respectively. No changes in fatal and non-fatal overdose rates were reported by 32% (n=24) of the countries (Figure 5, Table 2). Details of changes in mortalities and overdose rates are reported in supplementary materials.

**Figure 5.**
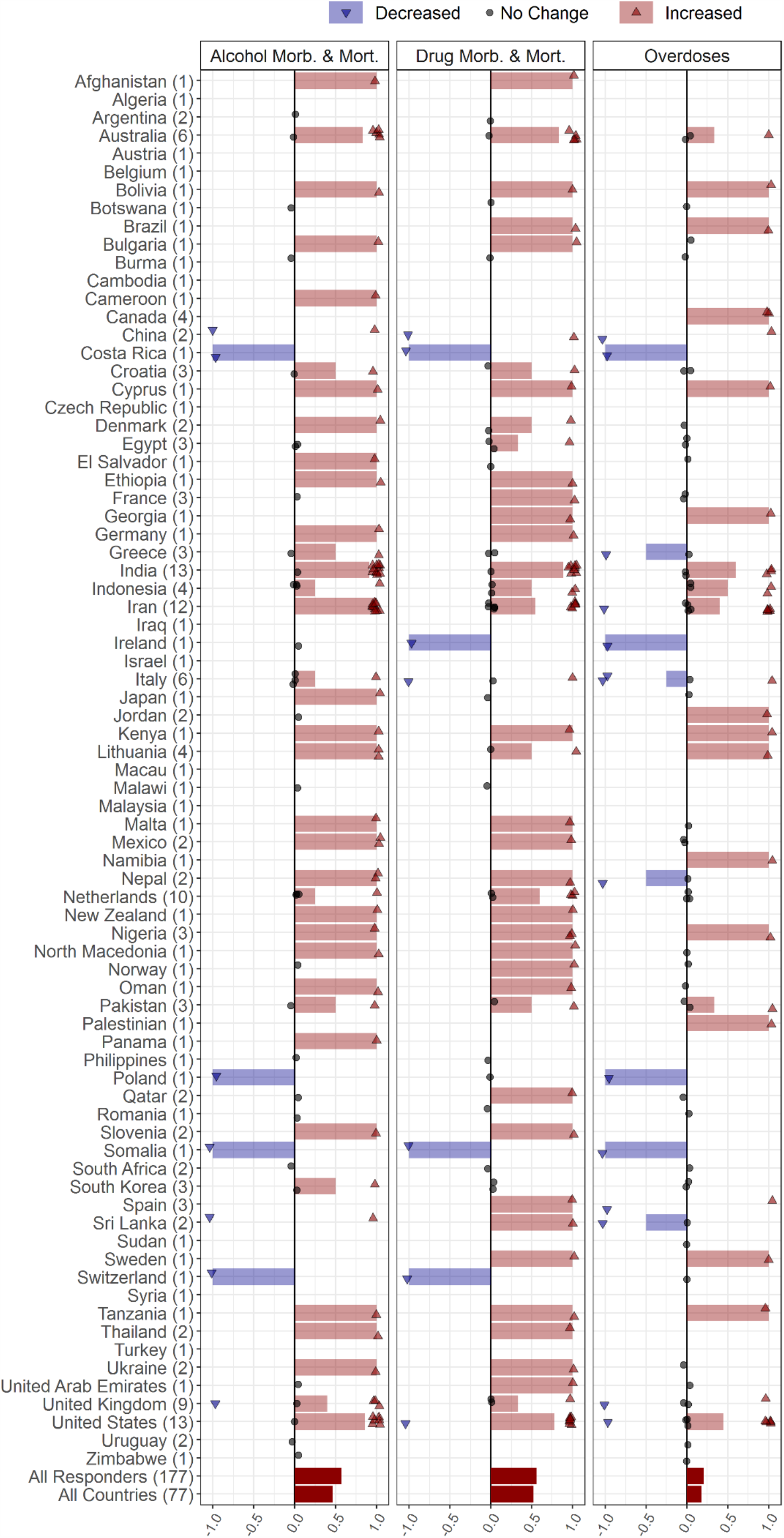
Changes in mortality, morbidity and overdose in people with SUD during the COVID-19 pandemic reported by 177 responders from 77 countries around the world. Responders were asked to report changes in morbidity or mortality rates in people with SUD and changes in fatal and non-fatal overdose episodes through the following options: *Increased, Decreased, I do not know, I do not like to answer, Not applicable*. Countries’ names are sorted in alphabetical order and the number of each countries’ responders are mentioned in front of the names. Each response is indicated as a single dot for *no change* or up and down triangles for *increased* and *decreased* answers respectively, with a minor jitter for better visualization. The reported answers are represented as -1 for *decreased*, 1 for *increased* and 0 for *no change; I do not know, I do not like to answer* and *Not applicable* answers are not shown in the figure. The mean of all responses, regardless of their originated countries and without considering those who didn’t know the answer, alongside the average answers of all countries, regardless of the number of responders in each country, are addressed in the last two rows below the countries’ names. (SUD: Substance Use Disorder).

### 3.7. Risky behaviors

Responders provided information about changes in risky behaviors among people with SUDs in their countries during the COVID-19 pandemic (figure 6, supplementary table 1). Information related to risky behaviors consisted of increased/switching to injection, sharing drug use equipment, needle and syringe sharing, and risky sexual behaviors. Sixteen percent (n=29) of the responders reported that injection among people with SUDs has increased, while 33% (n=58) reported no change in numbers of people injecting drugs or people switching to injection. Fifty-one percent (n=90) chose the ‘others’ option indicating lack of information or reluctance in responding to this question. Twenty-three percent (n=41) of the responders reported that sharing drug use equipment (i.e., paraphernalia) has increased, while 25% (n=44) reported no change. Fifty-two percent (n=92) chose the ‘others’ option indicating lack of information or reluctance in responding. Twenty-one percent (n=38) reported that sharing needle and syringe has increased, while 24% (n=43) reported no change. Fifty-four percent (n=96) chose the ‘others’ option indicating lack of information or reluctance in responding to this question. Twenty-three percent (n=41) reported that risky sexual behaviors have increased, while 22% (n=39) reported no change. Fifty-five percent (n=97) chose the ‘others’ option. Responders reported an increase in the behavioral addictions during the pandemic (Supplementary Figure 1).

**Figure 6.**
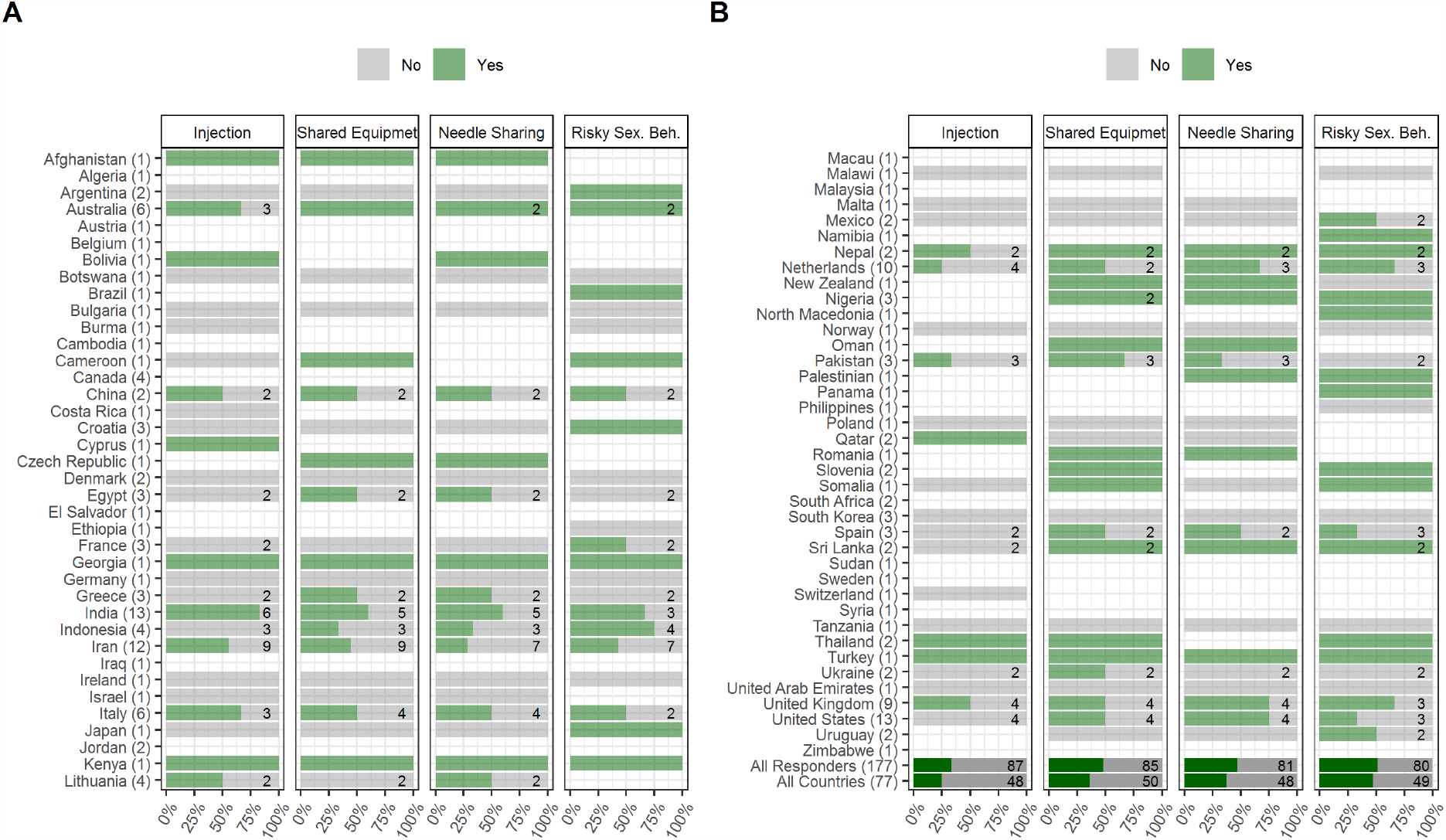
Changes in risky behaviors including shifting to injection, using shared drug use equipment, needle sharing and risky sexual behaviors during the COVID-19 pandemic period, reported by 177 responders from 77 countries globally. Responders were asked to report changes in risky behaviors (injection, shared drug use equipment, needle sharing and risky sexual behaviors) through the following options: *Yes, No, I do not know, I do not like to answer, Not applicable*. Countries’ names are sorted in alphabetical order and the number of each countries’ responders are mentioned in front of the names. Numbers of responders who reported *Yes* or *No* answers to each question are demonstrated inside the bars (If nothing is written, it indicates that there was only one response within *Yes* and *No* answers). The percentages shown by the bars are also based on only *Yes* or *No* answers. The mean percentages of all responses, regardless of their originated countries and without considering those who reported other than *Yes* and *No* answers, alongside the mean percentage answers of all countries, regardless of the number of responders in each country, are addressed in the last two rows below the countries’ names. (Risky Sex. Beh.: Risky Sexual Behaviors,).

### 3.8. COVID-19 overall impact on SUDs

Responders provided an overall rating of the general impact of the COVID-19 pandemic on people with SUDs in their countries (Figure 7). Oman, Kenya, and Georgia rated the highest severity of COVID-19 impact on people with SUDs (ratings of 10/10), while Botswana and Afghanistan rated the lowest severity for this impact in their countries (ratings of 2/10).

**Figure 7.**
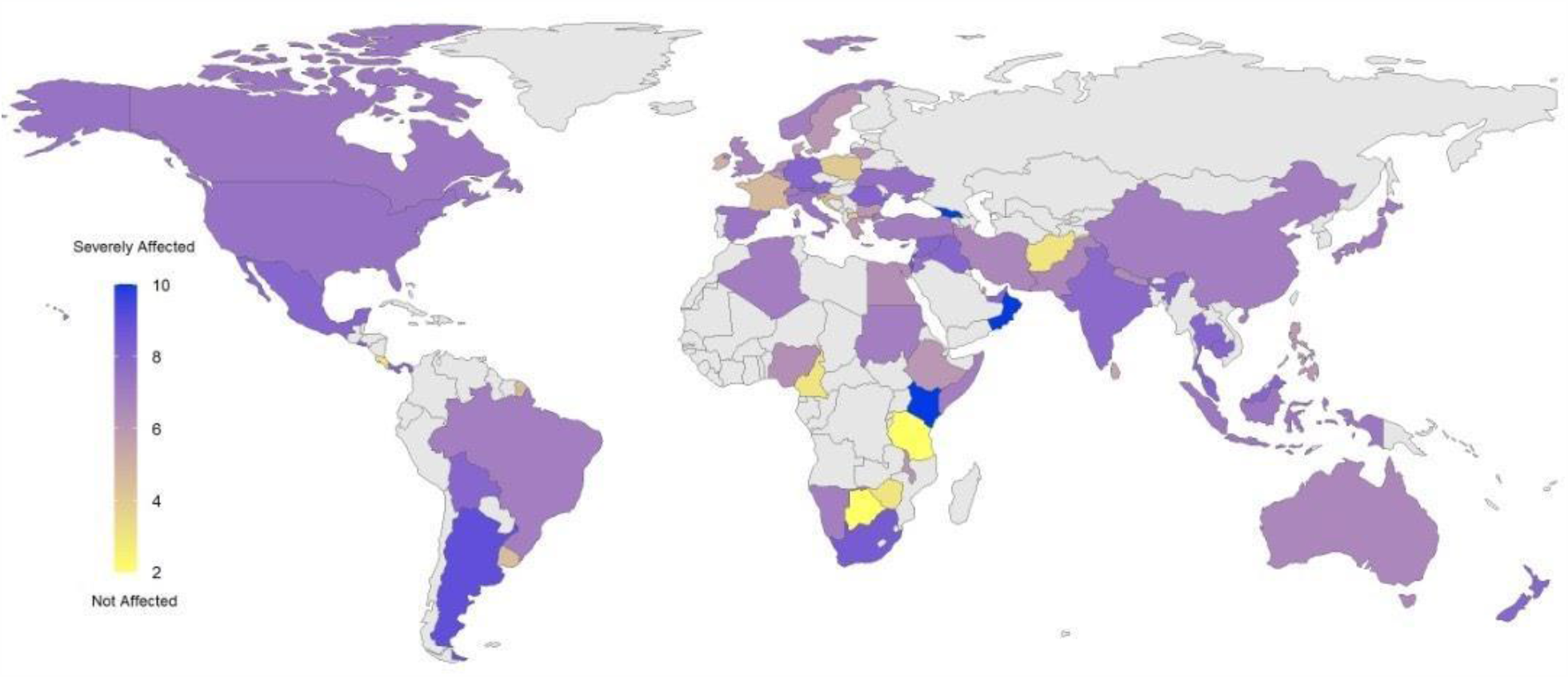
Severity of being affected by COVID-19 outbreak among people with SUDs globally. Addiction medicine professionals were asked to report how seriously people with SUDs in their countries have been affected by the COVID-19 pandemic using a range of between 1 to 10; 1 representing *Not affected*, demonstrated with yellow in the beginning of the spectrum and 10, representing *Severely affected* at the end of the spectrum, indicated with blue. Responses are collected from April 4^th^, 2020 during a 5-week period.

## 4. Discussion

According to the results of this first ever COVID-19 and SUD global survey with the contribution of 177 addiction medicine professionals/policy makers from 77 countries, the majority of responders believed that in their countries, people with SUDs had been seriously affected by the COVID-19 outbreak. They mostly believed that prices for alcohol and drugs have risen and they have become less available during the pandemic. In regard with alterations in use patterns, responders perceived an increase in use of alcohol, cannabis, prescribed opioids, and sedative/hypnotics use, and a decrease in the use of amphetamines, cocaine, and opiates. Most responders reported increases in complications related to drug and alcohol use including increased morbidity and mortality in people with SUDs.

Alterations in levels of alcohol consumption during pandemic is similar to those reported during prior social crises, like the 2008-2009 economic downturn (19). Changes in alcohol consumption may arise from two potentially contradictory, however interacting mechanisms: (1) a problematic increase, usually stemming from distress that is being experienced especially at the beginning of a crisis, or in an attempt to ‘stockpile’; or (2) a decrease due to the lack of access and financial difficulties which may lead to withdrawal (20). Current reports from Australia indicate increases in purchases of alcoholic beverages during lockdown potentially due to the first mechanism (21). However, India seems to be encountering a surge in numbers of individuals withdrawing from alcohol (5, 22). These independent reports from Australia and India are in line with our survey findings (figure 2). Initial reports from Australia and the United States indicate overall increases in alcohol sales, especially in online alcohol delivery subsectors (21), although specific data from industry on alcohol supply are largely lacking.

However, there was no consensus among our survey responders on alcohol supply as one third each reported increase, decrease or no change. Approximately half of our survey responders believed that there is no change in alcohol cost during the pandemic. This is while almost another half reported an increase in alcohol prices. We couldn’t find any relevant reports indicating alcohol price alterations. Further data are needed as the pandemic progresses and hopefully resolves.

There are currently concerns about morbidity and mortality spikes within people with AUDs and alcohol-associated liver disease (ALD) during the pandemic (23). The surveys’ results support the idea that these spikes can be seen among people with AUDs. Reports from Iran describe methanol poisoning of around 5,000 people with nearly 700 deaths, which may be due to lack of education and illegal and uncontrolled alcohol sales because of alcohol bans in Iran (10, 24, 25). However, to the best of our knowledge, there are yet no specific reports demonstrating the extent of alcohol overdose. The same pattern also applies for drug-related mortalities and morbidities.

Survey results suggest increases in cannabis use in more than half of participating countries. The European Monitoring Centre for Drugs and Drug Addiction (EMCDDA) has investigated this matter through three large darknet markets (26) in the first three months of 2020 and reported overall increased market activity, mostly in relation to cannabis products (13, 27). This might show the initial effects of the pandemic in the European countries market, particularly before peaks in number of people infected by COVID-19 and subsequent widespread lockdowns.

Opiates, amphetamines and cocaine were generally reported to have a decrease or no change in patterns of usage in most countries. During the 2008 global financial crisis, drug use patterns were differentially impacted, with expenditures of money for drugs down 2 to 44 percent, termed as the “Great Recession” of drug use (19). Although there are preliminary reports suggesting that opioid use is a risk factor for ICU admission in H1N1 infections and a possible risk factor for mortality following COVID-19 infection, rumors about protective effects of opium use in Iran may have led to increased consumption (28, 29). In the US, an already severe opioid overdose crisis worsened since the COVID-19 pandemic, with 30 out of 50 states reporting increases in overdoses between March and June of 2020, with an increase in high potency synthetic opioids such as fentanyl in street supplies and decreased access to harm reduction and OUD treatment services cited as possible drivers of overdose increase (30-32). While concerns have been also raised regarding probable effects of substances on COVID-19 patients (4, 33, 34), more research is need on changes in drug use patterns and impacts on SUDs.

More than 80% of the countries reported increased use of sedatives and hypnotics. This rise in the demand for sedatives/hypnotics may be related to stressful situation of the COVID-19 pandemic and its consequences. Survey results also suggest increased use of prescription opioids, perhaps for similar reasons, and changes in services may be needed (35, 36). Canada, Australia, United Kingdom and Scotland facilitated pharmacy-based methadone-dispensing programs as prescribing opioid-related medications increased (36). This model may help managing withdrawal syndromes during lockdown-related periods. In the United States, rapid changes in policies provided support to facilitate service delivery for people in opioid treatment programs, such as larger quantities of dispensed methadone and buprenorphine and relaxed regulations around virtual prescriber visits to initiate and continue medications for OUD in order to help patients access and maintain access to medications (35, 37).

The EMCDDA has reported recently increases in the drug demands in European markets (13). The EMCDDA has also noted that due to increases in retail prices of cannabis and cocaine, the localized supply shortages may exist during the pandemic (12). The UNODC has reported that across all regions globally, many countries have noted a general shortage of different drugs at retail level, mostly due to reduction in imports or strict lockdown rules, resulting in fewer personal interactions for drug sales (14). The UNODC has also noted a heterogeneous situation on bulk supply, both across drugs and across different countries (14). The UNODC preliminary data were gathered from governmental authorities and open sources (media and UNODC field officers) (14). Our results agree with multiple aspects of these reports of drug supplies.

The UNODC reported countries with strict rules on social distancing such as the Czech Republic, United Kingdom, Italy and Iran have been facing increased street drug prices due to lack of availability (14). Other reports from drug-producing countries suggest drug price decrements perhaps as a result of stockpiling of drugs (14). Subsequently, the EMCDDA, along with the UNODC, have both noted that COVID-19 restrictions have generally led to increases in drug prices, including cocaine, heroin, amphetamines and cannabis, at the level of street markets (13, 14). Our survey results support these preliminary data reported by the UNODC and EMCDDA.

Responders mostly reported increases in the behavioral addictions during the current pandemic, which may partly confirm the existing concerns on this matter (38, 39). Other small studies suggest increases in addictive behaviors (39-41). Some forms of gambling may have decreased due to financial uncertainties, occupational problems, cessation of sporting events, closure of casinos and other factors (40, 41). Discussing another addictive behavior, gaming has been represented to be a coping mechanism during the current stressful conditions (42). Accordingly, gaming has increased among college students in India, may who use gaming as an anti-stress mechanism (42). Increased gaming has been occurring globally during the pandemic (43), as has been pornography viewing (44). These and other concerns have led to guidance about internet use during the pandemic (45).

ISAM conducted the first global survey in the field of addiction medicine and successfully sampled responses from 77 countries and 177 experts globally. This timely and rapid survey was designed in a multi-step fashion including literature review, expert communication, professional qualitative appraisal and finally a pilot study (18) and was able to rapidly and reliably address urgent gaps in knowledge during the current pandemic. However, there are limitations such as heterogeneity in numbers of responders from different countries and their disciplines and educational levels. The convenience sample also may impact response rates and other factors. The lack of validated measures is a limitation, as is the lack of options for open ended responses that would provide a window on the mechanisms driving reported trends. Given the dynamic nature of pandemics and lack of multicentric epidemiological studies, the survey is a timely approach to provide a snapshot of global clinical addiction medicine concerns during these unprecedented times.

The objective the ISAM survey was to provide initial, rapid preliminary evidence about how COVID-19 has affected different situational aspects experienced by people with SUDs globally in order to help reach a better understanding of the current status, provision of this information to international organizations and regional policy makers should help authorities plan for addressing urgent needs and providing suitable services not only in the current pandemic, but also in future similar situations. To properly respond to the emerging demands and situational shifts during the COVID-19 pandemic in the addiction treatment services across the world, at a macro (policy) level, it is critical to recognize the importance of (1) the social safety net and measures used to reduce the social inequality widening gap when such epidemics deteriorate an already vulnerable system, (2) responsive and publicly well-resourced healthcare with adequate supply of appropriate medication, (3) civil liberties which could help increased participation and a judicious response by law enforcement agencies, and (4) policies that have taken in justifying alcohol sales and cannabis dispensaries as essential services and legislation allowing pharmacists to provide maintenance medications such as benzodiazepines in order to guarantee safe supplies. At a meso (organizational) level, it is important that clinical experience and knowledge on localized drug supply, price and associated morbidities and mortality is shared within the organization in order to respond adequately. This makes it vital that organizations have a responsive continuity plan that can change with the needs of the population throughout the acute stage of the pandemic. It is also important to establish, support and sustain varied digital platforms to allow better access to treatment for drug and alcohol using populations and minimize morbidities and possibly mortality.

Establishing jointly advocacy groups of service users and providers is also critical. At a micro (individual) level, it is important to (1) establish a mechanism for shared decision making through effective communication channels, (2) build the therapeutic environment that welcomes and encourages participation of peer, third sector and/or frontline workers who are also involved in the care of the individuals in care, (3) support a psychologically informed environments and interventions considering stress, uncertainties, isolation, and mental health, and (4) consider providing harm minimization and/or public protection messages and equipment to all in care and others.

In this unique global survey, experts in addiction medicine provided information on changes in regional alcohol and drug availability, price, usage and related complications. Reported decreases in alcohol and drug supplies appear partly attributable to lockdowns, import/export limitations and strict regulations. Reduced availability may have generated increases in prices. Reported increases in use of alcohol, cannabis, prescribed opioids, and sedative/hypnotics use may reflect their legal availability (in online markets, drugstores and dispensaries) while decreased use of amphetamines, cocaine, and opiates may related to decreased availability due to social distancing, lockdown regulations and increased prices. Changed drug use patterns may not only impact people with SUDs, but also give rise to risky behaviors and related complications. Most issues may potentially be preventable if future lockdown regulations are accompanied by enhanced service provision for at-risk communities.

## Data Availability

Survey data is available upon request

## Supplementary materials

### Supplementary Methods

**Supplementary Method 1. Responders’ global distribution**. Overall, 177 responders from 77 countries participated in the survey. The countries that provided information for this survey consisted of Afghanistan, Algeria, Argentina, Australia, Austria, Belgium, Bolivia, Botswana, Brazil, Bulgaria, Burma, Cambodia, Cameroon, Canada, China, Costa Rica, Croatia, Cyprus, Czech Republic, Denmark, Egypt, El Salvador, Ethiopia, France, Georgia, Germany, Greece, India, Indonesia, Iran, Iraq, Ireland, Israel, Italy, Japan, Jordan, Kenya, Lithuania, Macau, Malawi, Malaysia, Malta, Mexico, Namibia, Nepal, Netherlands, New Zealand, Nigeria, North Macedonia, Norway, Oman, Pakistan, Palestine, Panama, Philippines, Poland, Qatar, Romania, Slovenia, Somalia, South Africa, South Korea, Spain, Sri Lanka, Sudan, Sweden, Switzerland, Syria, Tanzania, Thailand, Turkey, Ukraine, United Arab Emirates, United Kingdom, United States, Uruguay, Zimbabwe.

**Supplementary Method 2. Survey questions related to the situational assessment**. The situational assessment section composed of the following questions: “Which of these substances went through changes in terms of usage pattern in your country?” (Increased/ decreased/ no changes), “Which of these substances went through changes in terms of price in your country?” (Increased/ decreased/ no changes), “Which of these substances went through changes in terms of supply in your country?” (Increased/ decreased/ no changes). The questioned substances included alcoholic beverages, cannabis (i.e. marijuana and synthetic cannabinoids such as spice, K2, etc.), opiates (i.e. opium, heroin, opium residue, etc.), amphetamine type stimulants (i.e. amphetamine, methamphetamine, MDMA, etc.), cocaine (including crack cocaine), sedative and hypnotics (i.e. Benzodiazepines, Barbiturates, etc.), and prescription opioids (e.g., Oxycodone, Oxycontin, Tramadol, Diphenoxylate, etc.). Other questions were “Have morbidities or mortalities increased in people with alcohol use disorders (AUDs) during the period of this pandemic?” (Severe, slight, no change in morbidities or mortalities), “Have morbidities or mortalities increased in people with SUDs during the period of this pandemic?” (Severe, slight, no change in morbidities or mortalities), “Have fatal and non-fatal overdose episodes changed among people with SUDs during the period of this pandemic?” (Severe, slight, no change in overdoses), “Have there been any changes in risky behaviors among people with SUDs during the period of this pandemic?” (risky behaviors consist of increased/switch to injection, sharing drug use equipment, needle and syringe sharing, and risky sexual behaviors), and “In general, how serious do you think people with SUDs in your country are affected by the COVID-19 outbreak?” (rate the severity on a 0-10 Likert scale).

### Supplementary Results

#### Drug use changes

Over 63% (n=49) of the countries reported that alcohol use increased, whereas 25% (n=19) and 8% (n=6) reported a decrement and no change in alcohol use pattern, respectively. Approximately 4% (n=3) responded that they do not know or are unwilling to answer this question. Approximately 42% of the countries (n=32) reported that cannabis use increased, whereas 26 % (n=20) and 22% (n=17) reported a decrease and no change in cannabis use pattern, respectively. Approximately 10% (n=8) responded that they do not know or are unwilling to answer this question. Approximately 18% (n=14) of the reported that opiates use increased, whereas 31% (n=24) and 20% (n=16) reported a decrement and no change in opiates use pattern, respectively. Approximately 30% (n=23) responded that they do not know or are unwilling to answer this question. Approximately 18% (n=14) of the countries reported that amphetamines use increased, whereas 29% (n=22) and 20% (n=15) reported a decrement and no change in amphetamines use pattern, respectively. Approximately 33% (n=26) responded that they do not know or are unwilling to answer this question. Approximately 14% (n=10) of the countries reported that cocaine use increased, whereas 29% (n=23) and 19% (n=15) reported a decrement and no change in cocaine use pattern, respectively 38% (n=29) responded that they do not know or are unwilling to answer this question. Approximately 64% (n=50) of the reported that sedatives and hypnotics use increased, whereas 6% (n=5) and 11% (n=9) reported a decrement and no change in sedatives and hypnotics use pattern, respectively. Approximately 18% (n=14) responded that they do not know or are unwilling to answer this question. Approximately 41% (n=32) of the countries reported that prescription opioids use was increased, whereas 11% (n=8) and 21% (n=16) reported a decrement and no change in prescription opioids use pattern, respectively. Approximately 27% (n=21) responded that they do not know or are unwilling to answer this question.

#### Drug supply changes

Over 31% (n=24) of the countries reported that the alcohol supply increased, whereas 34% (n=26) and 28% (n=21) reported a decrement and no change in the alcohol supply pattern, respectively.

Approximately 7% (n=6) responded that they do not know or are unwilling to answer this question. 20% (n=15) of the countries reported that the cannabis supply increased, whereas 37 % (n=29) and 24% (n=18) reported a decrement and no change in the cannabis supply pattern, respectively. Approximately 19% (n=15) responded that they do not know or are unwilling to answer this question. Only 8% (n=6) of the countries reported that opiates supply increased, whereas 41% (n=31) and 18% (n=14) reported a decrement and no change in opiates supply pattern, respectively. Approximately 33% (n=26) responded that they do not know or are unwilling to answer this question. Only 9% (n=7) of the countries reported that amphetamines supply increased, whereas 38% (n=29) and 18% (n=14) reported a decrement and no change in amphetamines supply pattern, respectively. Approximately 35% (n=27) responded that they do not know or are unwilling to answer this question. Only 9% (n=7) of the countries reported that the cocaine supply was increased, whereas 34% (n=24) and 18% (n=14) reported a decrement and no change in cocaine supply pattern, respectively. Approximately 39% (n=30) responded that they do not know or are unwilling to answer this question.

#### Drug price changes

Over 29% (n=23) of the countries reported that alcohol price increased, whereas 4% (n=3) and 54% (n=42) reported a decrement and no change in alcohol price, respectively. Approximately 13% (n=10) responded that they do not know or are unwilling to answer this question. 39% (n=30) of the countries reported that cannabis price increased, whereas 3% (n=2) and 30% (n=23) reported a decrease and no change in cannabis price, respectively. Approximately 28% (n=22) responded that they do not know or are unwilling to answer this question. 37% (n=29) of the countries reported that opiates price increased, whereas 2% (n=2) and 18% (n=14) reported a decrement and no change in opiates price, respectively. Approximately 43% (n=33) responded that they do not know or are unwilling to answer this question. 28% (n=21) of the countries reported that amphetamines price increased, whereas 0% (n=0) and 23% (n=18) reported a decrement and no change in amphetamines price, respectively. Approximately 49% (n=37) responded that they do not know or are unwilling to answer this question. Approximately 43% (n=33) of the countries reported that cocaine price increased, whereas 3% (n=2) and 39% (n=30) reported a decrease and no change in cocaine price, respectively. Approximately 15% (n=12) responded that they do not know or are unwilling to answer this question.

#### Morbidities, mortalities and overdose rate changes

41% (n=72) of responders reported that morbidities and mortalities have increased among people with AUDs during this pandemic, while 4% (n=7) and 19% (n=34) reported a decrement and no change regarding this matter, respectively. 36% (n=64) responded that they do not know or are unwilling to answer this question. Approximately 38% (n=68) of the reported that morbidities and mortalities have increased among people with SUD during this pandemic, while 4% (n=7) and 19% (n=34) reported a decrement and no change regarding this matter, respectively. Approximately 38% (n=68) responded that they do not know or are unwilling to answer this question. Approximately 20% (n=35) of the responders reported that fatal and non-fatal overdoses have increased among people with SUD, while 8% (n=14) and 30% (n=53) reported a decrement and no change in overdoses, respectively. Approximately 42% (n=75) responded that they do not know or are unwilling to answer this question.

## Supplementary Figures

**Supplementary Figure 1.**
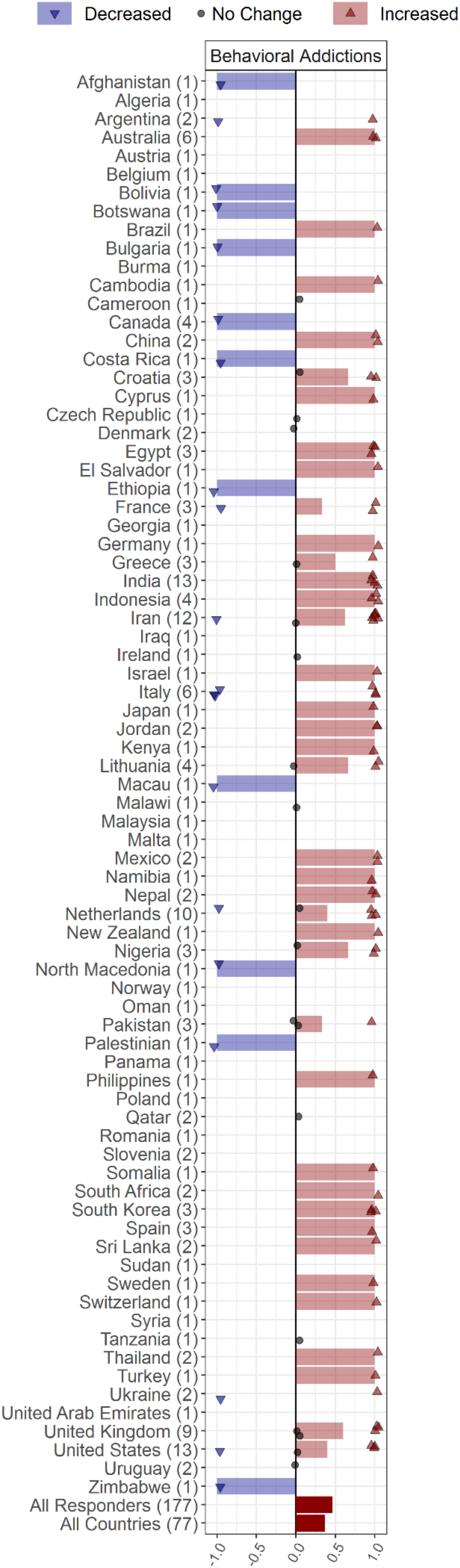
Changes in behavioral addictions including gambling and gaming during the COVID-19 pandemic, reported by 177 responders from 77 countries around the world. Responders were asked to report changes in behavioral addiction rates in their countries through the following options: *Increased, Decreased, No change, I do not know*. Countries’ names are sorted in alphabetical order, and the number of each countries’ responders is mentioned in front of the names. Each response is indicated as a single dot for *no change* or up and down triangles for *increased* and *decreased* answers, respectively, with a minor jitter for better visualization. The reported answers are represented as -1 for *decreased*, 1 for *increased*, and 0 for *no change; I do not know* answers are not shown in the figure. The mean of all responses, regardless of their originated countries and without considering those who didn’t know the answer, alongside the average answers of all countries, regardless of the number of responders in each country, are addressed in the last two rows below the countries’ names. The rates of behavioral addictions in the countries and responders that responded to this question have been increased by 40% and 50%, respectively. Approximately 86% (n=66) of the countries reported that behavioral addictions rates had increased, whereas 14% (n=11) of the countries reported that behavioral addictions rates had decreased in their countries during the COVID-19 pandemic.

## Supplementary Tables

**Supplementary Table 1.** Information related to risky behaviors including injection, sharing drug use equipment, needle and syringe sharing, and risky sexual behaviors among responders and countries.

|  | Responders (177) |  |  |  |  |  | Countries (77) |  |  |  |  |  |
| --- | --- | --- | --- | --- | --- | --- | --- | --- | --- | --- | --- | --- |
|  | No |  | Yes |  | Others |  | No |  | Yes |  | Others |  |
|  | N | % | N | % | N | % | N | % | N | % | N | % |
| Injection | **5** | **33** | 2 | 16 | 9 | 51 | **2** | **36** | 1 | 13 | 3 | 50 |
|  | **8** | **%** | 9 | **%** | 0 | **%** | **8** | **%** | 0 | **%** | 9 | **%** |
| Shared Drug Use Equipment | **4** | **25** | 4 | 23 | 9 | 52 | **2** | **26** | 1 | 24 | 3 | 50 |
|  | **4** | **%** | 1 | **%** | 2 | **%** | **0** | **%** | 8 | **%** | 9 | **%** |
| Needle Sharing | **4** | **24** | 3 | 21 | 9 | 54 | **2** | **27** | 1 | 21 | 4 | 52 |
|  | **3** | **%** | 8 | **%** | 6 | **%** | **1** | **%** | 6 | **%** | 0 | **%** |
| Risky Sexual Behaviors | 3 | 22 | **4** | **23** | 9 | 55 | **2** | **25** | 1 | 25 | 3 | 49 |
|  | 9 | **%** | **1** | **%** | 7 | **%** | **0** | **%** | 9 | **%** | 8 | **%** |

